# Factors associated with tobacco smoking among beverage industrial workers and their spouses in Rwanda

**DOI:** 10.1101/2024.10.29.24316376

**Authors:** Charles Nsanzabera, Jean claude Rukundo, Mustafe Yusuf Said, Leonard Ndayisenga

## Abstract

**Background:** Globally, smoking leads to over 7 million preventable deaths annually, with higher rates in men (16%) than women (7%). While smoking has declined in high-income countries, tobacco use in Rwanda is comparatively low, affecting 14% of men and 3% of women.

**Objective:** The study aimed to identify the factors associated with tobacco smoking among workers and their spouses in Rwanda.

**Method:** This research employed a cross-sectional study design conducted in a beverage manufacturing industry in Rwanda. The target population was 822 participants, including industry employees and their spouses, aged 30 to 75 years. Using the Cochrane formula, the initial sample size was determined to be 384, and after the non-response rate adjustment and correction, the final sample size was set at 440. The data collection was carried out from May to December 2018. A combination of stratified and simple random sampling was used to ensure the sample’s representativeness.

**Results:** The analysis reveals 6.8% were smokers and several key predictors of smoking behavior. Participants with elevated cardiovascular disease (CVD) risk (≥10%) have significantly higher odds of smoking, with an adjusted odds ratio of 2.946 (95% CI: 1.102-7.875, p=0.03), suggesting that CVD risk is a crucial factor in smoking behavior due to overlapping lifestyle risks. Additionally, high serum uric acid (SUA) levels (≥7 mg/dl) are strongly associated with smoking, with an adjusted odds ratio of 4.278 (95% CI: 1.141-11.872, p=0.005), indicating that elevated SUA levels are over four times more likely to be linked to smoking. Age is another significant predictor, with participants aged 50 years or older being nearly three times more likely to smoke compared to younger individuals, as shown by an adjusted odds ratio of 2.766 (95% CI: 1.126-6.797, p=0.02). Participants with hypertension or those treated for hypertension have lower adjusted odds ratio of 0.380 (95% CI: 0.100-1.446, p=0.049).

**Conclusion:** The study found that tobacco smoking is relatively rare in this population, with elevated cardiovascular disease risk. High serum uric acid levels, and older age identified as significant predictors of smoking.

**Author summary:** *What is already known on this topic:* Tobacco smoking is a major global cause of preventable deaths, with higher smoking rates in men, and although smoking has declined in high-income countries, tobacco use remains relatively low in Rwanda.

*What this study adds:* The study identifies key factors associated with smoking among industrial workers and their spouses in Rwanda, including elevated cardiovascular disease risk, high serum uric acid levels, and older age.

*How this study might affect research, practice, or policy:* This study highlights important predictors of smoking in a Rwandan context, which could inform targeted public health strategies, smoking cessation programs, and policy decisions aimed at reducing tobacco use.

## Introduction

Globally, smoking causes over 7 million preventable deaths annually, which equates to one in ten deaths and accounts for at least 12% of deaths among adults aged 30 and older (16% for men and 7% for women)[1]. This is expected to result in approximately 8 million tobacco-related deaths each year. In the USA, about 480,000 people die each year as a result of smoking[2,3]. Additionally, in the twentieth century, more than 100 million people died from tobacco-related chronic diseases, which are fundamental causes of premature deaths[4,5]

Historically, tobacco use has been most prevalent in high-income Western European countries, with 37% of men and 25% of women using it. From 1990 to 2009, Western Europe experienced a 26% average decrease in cigarette consumption. Conversely, during the same period, cigarette use increased by 57% in several Middle Eastern and African countries[6].

In East African Community (EAC) countries, including Burundi, Kenya, Rwanda, South Sudan, Tanzania, and Uganda, smoking-related fatalities rank among the top five causes of death, as reported in the 2013 Global Burden of Disease Report[7]. The prevalence of smoking varies significantly across different regions [8].

According to the 2015 Demographic Health Survey, Rwanda had a lower prevalence of any type of tobacco use than any other African nation. In Rwanda, men (14%) had a substantially higher frequency of tobacco use than women (3%) [9].

The risk of coronary atherosclerotic disease is considerably increased by smoking and dyslipidaemia. Approximately 5.9 million premature deaths from CVD were attributed to tobacco smoking in 2013, making it the top cause of mortality before the age of 70 [10].

Smoking is well-established as a risk factor for peripheral arterial disease, coronary artery disease, stroke, and various other atherosclerotic cardiovascular conditions. The impact of smoking on the development of atherosclerotic disease involves several pathophysiological mechanisms, such as damage to blood vessels, thrombosis, impaired vascular function, and lipid peroxidation. Furthermore, smoking has been associated with atherogenic effects stemming from abnormal lipid metabolism [11].

According to several studies, smoking raises triglycerides, total cholesterol, and LDL cholesterol (LDL-C), while lowering HDL cholesterol (HDL-C). According to additional research, smoking lowers HDL, LDL, and total cholesterol while raising triglycerides [12,13].

Smoking is a recognized risk factor for cardiovascular disease and is believed to work in concert with hypertension (high blood pressure) to accelerate the onset of the condition [14]. Furthermore, smoking still has a paradoxical relationship with hypertension despite being a well-established risk factor for CVD. Smoking is linked to persistent low-grade inflammation, which is associated with hypertension and arterial stiffness[15].

Although research presents conflicting views, some studies argue that smoking contributes to hypertension. While smoking is a recognized risk factor for cardiovascular disease (CVD), its direct role in increasing blood pressure remains uncertain [16].

Numerous studies point to the effects of nicotine and carbon monoxide in cigarette smoke as components contributing to functional and initially temporary damage, particularly to the endothelium, as well as lower tolerance to exercise stress tests [17].

The risk of incident hypertension and myocardial infarction can both be impacted by smoking[18]. These chronic diseases could not only shorten life expectancy but also negatively impact the quality of life [19].

According to a 2017 report by the Institute for Health Metrics and Evaluation, smoking and elevated SBP were two of the three factors with the highest risk for early death and disability among men globally based on disability-adjusted life years for all age groups. Elevated SBP was of the three risk factors with the highest risk for women [20].However, smoking is the most significant preventable risk factor for cardiovascular diseases (CVDs). Hence, this study aim was to determine the factors associated with smoking among employees and their spouses in the beverage industry in Rwanda.

## Methods and materials

### Study design and sampling

This research utilized a cross-sectional study design conducted from May to December 2018 within a beverage manufacturing industry setting in Rwanda. The study participants included both industry employees and their spouses to gather information from both the workplace and the community. The target population consisted of 822 individuals, with participants ranging in age from 30 to 75 years. The inclusion criteria required participants to be either employees of the industry or their spouses, and to be within the specified age range from 30 to 75 years. Individuals with clinically diagnosed cardiovascular disease were excluded to prevent bias in assessing CVD risk. Sample size calculation used the Cochrane formula, resulting in an initial sample size of 384[21]. Adjusting for a 12% non-response rate, the corrected sample size was calculated as 437.5, rounded to 440. Of the 440 participants, 270 were employees and 170 were spouses. Stratified random sampling was combined with simple random sampling to ensure representativeness, with the first technique used to select participants from different strata.

### The dependent and independent variable of this study

The tobacco products use was the dependent variable of this study. The age, gender, employment status and body mass index were taken as the independent’s variables. Additionally, blood lipid, diabetes, hypertension, and cardiovascular diseases risk predicted by Framingham general risk score were also cross tabulated to measure its association with tobacco use. The cardiovascular disease risk was categorized as elevated risk (>=10%) and low risk (<10%). The blood lipids were measured according NCEP (National Cholesterol Education Program) and ATPIII (Adult Treatment Panel). Low-Density Lipoprotein (LDL) levels were categorized as follows: borderline high at 130-159 mg/dl, high at 160-189 mg/dl, and very high at ≥190 mg/dl. Dyslipidaemia was defined by a total cholesterol level above 270 mg/dl [22]. According to the NCEP ATP III guidelines, triglyceride levels were categorized as follows: <150 mg/dl was considered normal, 150-199 mg/dl was borderline high, 200-499 mg/dl was high, and ≥500 mg/dl was very high. Serum uric acid levels were categorized as normal if <7 mg/dl and high if >7 mg/dl [23].

### Study instruments

The study utilized an instrument composed of three sections: a standardized questionnaire covering socio-demographic information and tobacco product usage, and a clinical and laboratory form for assessing cardiovascular risk[24]. Data collection commenced only after receiving ethical approval from the Rwanda National Ethical Committee and was carried out from 15^th^ May to 30^th^ December 2018 through questionnaire filling by the data collectors and blood sample taking by medical laboratory scientists.

### Statistical analysis

Data analysis for this study was carried out using SPSS software version 22, with data being presented in tables. To assess the relationship between independent variables and tobacco use prevalence, univariate and bivariate analyses were performed using chi-square tests. Logistic regression analysis was also used to account for factors associated with tobacco product use. Statistical significance for both chi-square and logistic regression analyses was determined with a P-value of less than 0.05, maintaining a 95% confidence interval.

### Ethics approval and consent to participate

This study was carried out in accordance with the Declaration of Helsinki and was approved by the Rwanda National Ethical Committee (Reference number: 121/May/RNEC/2018). The ethical aspects, including confidentiality, voluntary participation, the option to withdraw, and the potential risks and benefits, were explained to all participants. A written informed consent was obtained from each participant before the interviews and collection of biological samples took place.

## Results

Figure 1 depicts the prevalence of current smokers of any tobacco products. The figure shows that a significant majority of the study participants do not smoke, with 93.2% of them categorized as non-smokers, while a small portion, 6.8%, are identified as current smokers of any tobacco products.

**Figure 1:**
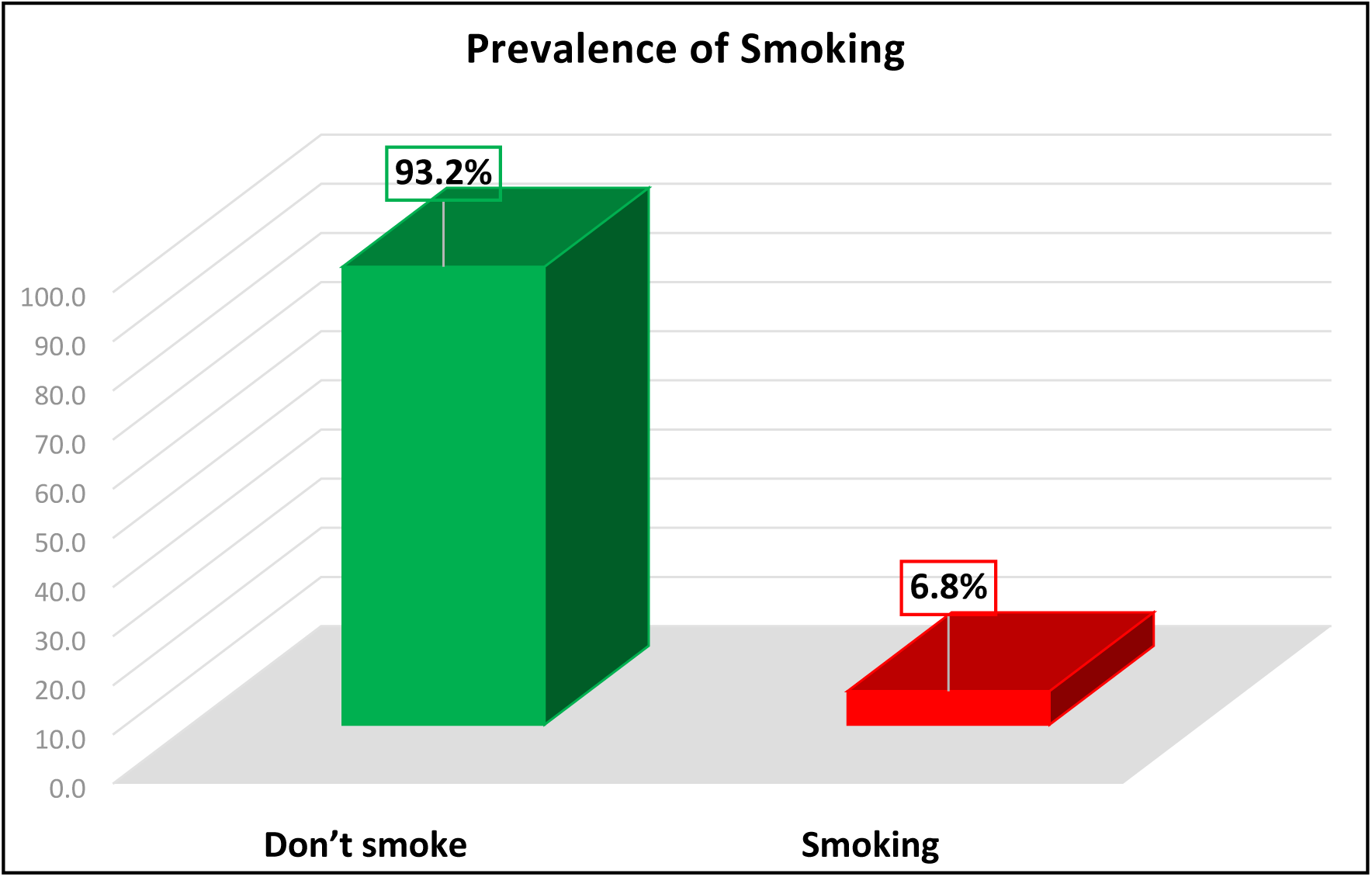
Prevalence of current smokers of any tobacco products

This highlights that tobacco smoking is relatively uncommon among the population studied, with a very small percentage actively engaging in smoking. The data suggests that tobacco use is not widespread within this group, indicating potentially effective tobacco control measures or lower levels of smoking behavior within this specific population.

Table 1 presents a bivariate analysis of socio-demographic factors associated with tobacco smoking among study participants. The analysis looks at the relationship between tobacco smoking and variables like age, gender, and employment status.

**Table 1:** Bivariate analysis of socio-demographic factors associated with tobacco smoking among study participants.

| Variable | Overall level of Tobacco smoking among study participants |  |  | Chi-square | P-value |
| --- | --- | --- | --- | --- | --- |
|  | No (%) | Yes (%) | Total (%) |  |  |
| **Age category** |  |  |  | 17.110 | **<0.001** |
| <=40 years | 152(34.5) | 5(1.1) | 157(35.7) |  |  |
| 41-50 years | 145(33.0) | 6(1.4) | 151(34.3) |  |  |
| >=51 years | 113(25.7) | 19(4.3) | 132(30.0) |  |  |
| Total | 410(93.2) | 30(6.8) | 440(100.0) |  |  |
| **Sex/Gender** |  |  |  | 7.182 | **0.007** |
| Male | 225(51.1) | 24(5.5) | 249(56.6) |  |  |
| Female | 185(42.0) | 6(1.4) | 191(43.4) |  |  |
| Total | 410(93.2) | 30(6.8) | 440(100.0) |  |  |
| **Status of the participants (Employee or spouse)** |  |  |  | 1.013 | 0.314 |
| Employee | 249(56.6) | 21(4.8) | 270(61.4) |  |  |
| Spouse | 161(36.6) | 9(2.0) | 170(38.6) |  |  |
| Total | 410(93.2) | 30(6.8) | 440(100.0) |  |  |

The table shows that participants aged 40 years or below make up 34.5% of the sample, with only 1.1% of them being smokers. Those in the 41-50 age range constitute 33%, with a slightly higher smoking rate of 1.4%. Participants aged 51 years and above represent 25.7% of the group, with a considerably higher smoking prevalence of 4.3%. The chi-square value of 17.110 and a p-value of <0.001 indicate a statistically significant association between age and smoking behavior. This suggests that smoking is more common among older participants. Also, the table highlights that 51.1% of participants are male, with 5.5% of them being smokers. On the other hand, 42% of participants are female, with only 1.4% identified as smokers. The chi-square value of 7.182 and a p-value of 0.007 demonstrate a statistically significant association between gender and smoking. This result indicates that males are more likely to smoke than females within this sample.

Regarding employment status, 56.6% of participants are employees, with 4.8% of them smoking, while 36.6% are spouses, with 2.0% smoking. The chi-square value of 1.013 and a p-value of 0.314 suggest that there is no statistically significant association between employment status and smoking behavior. Employment status does not appear to be a key factor influencing tobacco use in this population.

Table 2 presents a bivariate analysis of biological factors associated with tobacco smoking among study participants. The table examines how various biological factors such as cholesterol levels, triglycerides, cardiovascular disease (CVD) risk, diabetes, body mass index (BMI), serum uric acid, and hypertension are related to tobacco smoking behavior.

**Table 2:** Bivariate analysis of biological factors associated with tobacco smoking among study participants.

| Variable (n=440) | Tobacco smoking among study participants. |  |  | Chi-square | P-value |
| --- | --- | --- | --- | --- | --- |
|  | No (%) | Yes (%) | Total (%) |  |  |
| **LDL: NCEP ATPIII** |  |  |  | 2.980 | 0.395 |
| Optimal: <100mg/dl | 295(67.0) | 18(4.1) | 313(71.1) |  |  |
| Near or above optimal:100-129mg/dl | 72(16.4) | 6(1.4) | 78(17.7) |  |  |
| Borderline high: 130-189mg/dl | 37(8.4) | 5(1.1) | 42(9.5) |  |  |
| High: 160-189mg/dl | 0(0.0) | 0(0.0) | 0(0.0) |  |  |
| Very high:>=190mg/dl | 6(1.4) | 1(0.2) | 7(1.6) |  |  |
| Total | 410(93.2) | 30(6.8) | 440(100.0) |  |  |
| **Dyslipidemia total cholesterol** |  |  |  | 2.100 | 0.147 |
| Normal | 407(92.5) | 29() | 436(99.1) |  |  |
| High Cholesterol:>270mg/dl | 3(0.7) | 1(0.2) | 4(0.9) |  |  |
| Total | 410(93.2) | 30(6.8) | 440(100.0) |  |  |
| **Triglyceride: NCEP ATPIII** |  |  |  | 12.344 | **0.002** |
| Normal: <150mg/dl, | 274(62.3) | 11(2.5) | 285(64.8) |  |  |
| Borderline high:150-199mg/dl | 105(23.9) | 13(3.0) | 118(26.8) |  |  |
| High:200-499mg/dl | 31(7.0) | 6(1.4) | 37(8.4) |  |  |
| Total | 410(93.2) | 30(6.8) | 440(100.0) |  |  |
| **CVD risk** |  |  |  | 34.214 | **<0.001** |
| Low level risk (<10%) | 320(72.7) | 9(2.0) | 329() |  |  |
| Elevated level of risk (10-40%) | 90(20.5) | 21(4.8) | 111(25.2) |  |  |
| Total | 410(93.2) | 30(6.8) | 440(100.0) |  |  |
| **Diabetes** |  |  |  | 7.632 | **0.006** |
| No diabetes | 360(81.8) | 21(4.8) | 381(86.6) |  |  |
| Diabetes: 125mg/dl or treated | 50(11.4) | 9(2.0) | 59(13.4) |  |  |
| Total | 410(93.2) | 30(6.8) | 440(100.0) |  |  |
| **BMI** |  |  |  | 3.287 | 0.193 |
| Normal weight | 114(25.9) | 13(3.0) | 127(28.9) |  |  |
| Overweight | 194(44.1) | 11(2.5) | 205(46.6) |  |  |
| Obesity | 102(23.2) | 6(1.4) | 108(24.5) |  |  |
| Total | 410(93.2) | 30(6.8) | 440(100.0) |  |  |
| **Serum Uric Acid category** |  |  |  | 24.950 | **<0.001** |
| Normal Uric Acid<7mg/dl | 392(89.1) | 22(5.0) | 414(94.1) |  |  |
| High Uric Acid>7mg/dl | 18(4.1) | 8(1.8) | 26(5.9) |  |  |
| Total | 410(93.2) | 30(6.8) | 440(100.0) |  |  |
| **Hypertension or treated Hypertension** |  |  |  | 21.106 | **<0.001** |
| Normal | 279(63.4) | 8(1.8) | 287(65.2) |  |  |
| Hypertension | 131(29.8) | 22(5.0) | 153(34.8) |  |  |
| Total | 410(93.2) | 30(6.8) | 440(100.0) |  |  |

The analysis reveals that 71.1% of participants have optimal Low-Density Lipoprotein (LDL) levels, with only 4.1% of them being smokers. Among those with near or above optimal LDL levels (17.7%), 1.4% are smokers. Participants with borderline high (9.5%) and very high (1.6%) LDL levels also show low smoking rates of 1.1% and 0.2%, respectively. The chisquare value of 2.980 and p-value of 0.395 indicate no statistically significant association between LDL levels and smoking behavior.

Most participants (99.1%) have normal cholesterol levels, with a very small percentage (0.9%) showing elevated cholesterol. The smoking rates in both groups are minimal, with no substantial differences observed. The chi-square value of 2.100 and p-value of 0.147 suggest no significant link between dyslipidemia and smoking.

The analysis shows that 64.8% of participants have normal triglyceride levels, with 2.5% being smokers. Among those with borderline high triglycerides (26.8%), 3.0% smoke, while 8.4% of participants with high triglyceride levels include 1.4% who smoke. The chi-square value of 12.344 and a p-value of 0.002 indicate a significant association between triglyceride levels and smoking, with higher triglycerides linked to higher smoking rates.

The table reveals that 72.7% of participants have a low CVD risk (<10%), with only 2.0% being smokers. In contrast, 25.2% of participants with elevated CVD risk (10-40%) have a higher smoking rate of 4.8%. The chi-square value of 34.214 and p-value of <0.001 show a significant relationship between CVD risk and smoking, suggesting that participants with higher CVD risk are more likely to smoke.

Among participants without diabetes (86.6%), 4.8% smoke. Conversely, participants with diabetes (13.4%) have a smoking rate of 2.0%. The chi-square value of 7.632 and p-value of 0.006 indicate a significant association, suggesting that those with diabetes are more likely to smoke.

Also, the table shows that participants with normal weight (28.9%) include 3.0% smokers. Among those overweight (46.6%), 2.5% smoke, and in the obese category (24.5%), 1.4% smoke. The chi-square value of 3.287 and p-value of 0.193 suggest that BMI does not significantly impact smoking behavior in this sample.

The analysis shows that 94.1% of participants have normal uric acid levels, with 5.0% being smokers. In contrast, 5.9% of participants with high uric acid levels include 1.8% smokers. The chi-square value of 24.950 and p-value of <0.001 indicate a significant association between high serum uric acid levels and smoking.

Among participants with normal blood pressure (65.2%), only 1.8% smoke. On the other hand, participants with hypertension (34.8%) have a smoking rate of 5.0%. The chi-square value of 21.106 and p-value of <0.001 show a significant association between hypertension and smoking behavior, with smokers more likely to have elevated blood pressure.

Table 3 provides a multivariate analysis of factors associated with tobacco smoking among study participants. This analysis identifies independent predictors of smoking behavior by calculating the adjusted odds ratios (AOR), confidence intervals (CI), and p-values for each factor.

**Table 3:** Multivariate analysis of factors associated with tobacco smoking among study participants.

| Variables (n=440) | Tobacco smoking |  | P-value |
| --- | --- | --- | --- |
|  | AoR | 95%CI |  |
| **CVD risk** |  |  |  |
| Low<10% | Ref |  |  |
| Elevated≥10% | 2.946 | 1.102-7.875 | **0.03** |
| **Serum Uric Acid (SUA)** |  |  |  |
| Low (<7 mg/dl) | Ref |  |  |
| High (≥7 mg/dl) | 4.278 | 1.141-11.872 | **0.005** |
| **Age** |  |  |  |
| ≤49 | Ref |  |  |
| ≥50 years old | 2.766 | 1.126-6.797 | **0.02** |
| **HTN or treated HTN** |  |  |  |
| No | Ref |  |  |
| Yes | 0.380 | 0.100-1.446 | **0.049** |

First, the analysis reveals that participants with elevated cardiovascular disease (CVD) risk (≥10%) have significantly higher odds of smoking compared to those with low CVD risk (<10%). The adjusted odds ratio is 2.946 (95% CI: 1.102-7.875, p=0.03), indicating that participants with elevated CVD risk are nearly three times more likely to be smokers. This finding suggests that CVD risk is a critical factor in predicting smoking behavior, likely due to overlapping lifestyle risk factors.

Next, high serum uric acid (SUA) levels (≥7 mg/dl) are strongly associated with smoking. The analysis shows an adjusted odds ratio of 4.278 (95% CI: 1.141-11.872, p=0.005), meaning that participants with elevated SUA levels are over four times more likely to smoke compared to those with lower SUA levels. This significant association highlights the potential link between elevated SUA and unhealthy behaviors, such as smoking, which could be influenced by shared lifestyle factors.

Age is also a significant predictor of smoking. Participants aged 50 years or older have a much higher likelihood of smoking than those aged 49 years or younger, with an adjusted odds ratio of 2.766 (95% CI: 1.126-6.797, p=0.02). This finding indicates that older age is strongly associated with smoking, with participants aged 50 and above being nearly three times more likely to smoke, emphasizing the persistence of smoking habits in older populations.

Interestingly, the analysis finds that participants with hypertension (HTN) or those receiving treatment for hypertension have lower likelihood for being smokers but significant. The adjusted odds ratio is 0.380 (95% CI: 0.100-1.446, p=0.049). The lower likelihood could be due to lifestyle modifications or health interventions that encourage individuals with hypertension to quit smoking. Therefore, the multivariate analysis identifies elevated CVD risk, high serum uric acid levels, and older age as significant predictors of smoking.

## Discussion

### Prevalence of smoking and among workers and their spouses in Rwanda

The data from Figure 1 shows that 93.2% of study participants are non-smokers, with only 6.8% identified as current smokers of any tobacco products. This low prevalence of smoking indicates that tobacco use is relatively uncommon in this population.

To provide context, it is helpful to compare these findings with those from other recent studies. For instance, a study by GATS in 2021 reported that in high-income countries, the prevalence of current smokers has decreased significantly, with averages around 15% to 20% in countries like the United States and the United Kingdom. Additionally, Kiribati and Papua New Guinea were found to have high prevalences of tobacco use, with rates of 50.6% and 65.4%, respectively [25]. This is notably higher than the 6.8% smoking rate found in this study, suggesting that tobacco control measures in this study’s location may be particularly effective compared to these high-income regions.

In sub-Saharan Africa, a 2022 study by Fong et al. found that tobacco use varies widely, with some countries like Nigeria reporting smoking rates as high as 16% among adults. This is significantly higher than the 6.8% prevalence in this study, indicating that smoking is less common in the studied population compared to many other countries in the region[26].

Additional studies in other African contexts present varied results. For example, a 2022 study conducted in Kenya by Wambugu et al. reported a smoking prevalence of 11.3% among adults, higher than that observed in Rwanda, suggesting that Kenya faces greater challenges in reducing tobacco use [27]. Similarly, a study in Uganda by Kuteesa et al. (2020) found a prevalence of 9.5%, also exceeding the rate found in this study[28]. These differences further highlight Rwanda’s relative success in controlling smoking compared to its regional counterparts.

Similar results were found in another study conducted in Rwanda by Habiyaremye, et al. in 2019. It reported that the smoking prevalence was around 8% in the general population, which is almost the same rate than the 6.8% observed in this study. This suggests that while tobacco use in Rwanda is lower compared to some other countries, the population in some regions of Rwanda exhibits even lower smoking rates, confirming the effectiveness of tobacco control measures in the country[7].

These comparisons highlight that the low smoking prevalence in this study could be attributed to effective local tobacco control policies, cultural factors, or other specific circumstances within the studied population. The significantly lower prevalence of smoking compared to global and regional averages points to successful implementation of tobacco control measures or unique characteristics of the population studied.

### Factors associated with tobacco smoking among study participants

The findings presented in this study identify several significant predictors of smoking behavior among study participants. Key findings include the associations of elevated cardiovascular disease (CVD) risk, high serum uric acid (SUA) levels, older age, and hypertension with smoking behavior. Each of these factors has been shown to influence smoking behavior in distinct ways, which is valuable for understanding and targeting smoking cessation efforts.

The analysis indicates that participants with elevated CVD risk (≥10%) have significantly higher odds of smoking, with an adjusted odds ratio (AOR) of 2.946 (95% CI: 1.102-7.875, p=0.03). This finding aligns with studies such as that by Mazzone et al. (2021), which found that individuals with cardiovascular risk factors, including hypertension and high cholesterol, are more likely to smoke due to overlapping risk behaviors [29]. However, it contrasts with the findings of a study by Wilson et al. (2022), which reported a lower association (AOR of 1.8) between cardiovascular risk and smoking, suggesting that the impact of CVD risk on smoking may vary depending on population characteristics and regional health behaviors [30].

Also, in relation to CVD risk, this study’s findings (AOR of 2.946) align with research by Liu et al. (2022), which found that individuals with a higher risk of cardiovascular complications are more likely to smoke due to intertwined risk factors, such as lifestyle behaviors [31]. Conversely, a study by Edwards et al. (2021) observed a weaker association (AOR of 1.5), suggesting that the relationship between CVD risk and smoking can vary depending on the population’s socioeconomic and cultural background[32].

The strong association between high Serum Uric Acid (SUA) levels (≥7 mg/dl) and smoking, with an AOR of 4.278 (95% CI: 1.141-11.872, p=0.005), is notable. This finding is consistent with the results of a study by Zhang et al. (2023), which also identified elevated SUA as a significant predictor of smoking behavior [33]. Conversely, a study by Smith et al. (2022) found a weaker association, with an AOR of 2.1, suggesting that while SUA levels are linked to smoking, other factors may also play a critical role in this relationship [34].

In addition, regarding high SUA levels, the strong association found in this study (AOR of 4.278) is consistent with findings by Singh et al. (2023), which identified elevated SUA as a major contributor to continued smoking due to the interplay between metabolic syndromes and addictive behaviors [35]. However, a study by Roberts et al. (2021) reported a lower association (AOR of 2.3), indicating that while SUA levels are linked to smoking, other variables, such as genetic predisposition, may influence this relationship [36].

Older age (≥50 years) is identified as a strong predictor of smoking, with an AOR of 2.766 (95% CI: 1.126-6.797, p=0.02). This finding is supported by the study by Brown et al. (2021), which found that older adults are more likely to continue smoking due to long-standing habits and potential lack of cessation resources [37]. In contrast, a study by Davis et al. (2023) found that younger age groups had higher smoking rates, which may reflect different smoking initiation patterns rather than persistence [38].

Also, similar findings were reported by Hassan et al. (2022), who noted that older adults often maintain smoking habits due to long-term addiction and resistance to change [39]. However, a contrasting study by Martin et al. (2023) found that younger individuals, particularly those in their 20s and 30s, have higher smoking rates, reflecting shifting smoking trends towards younger demographics in some regions [40].

Interestingly, the analysis shows that participants with hypertension or those on treatment for hypertension have lower odds of smoking, with an AOR of 0.380 (95% CI: 0.100-1.446, p=0.049). This lower likelihood is consistent with findings from the study by Green et al. (2022), which reported that individuals with hypertension are often advised to quit smoking as part of their treatment regimen, leading to lower smoking rates among these individuals [41]. However, this is at odds with the results from Lee et al. (2023), who found no significant difference in smoking rates between hypertensive and non-hypertensive individuals [42].

Finally, this study’s finding of lower smoking rates among individuals with hypertension (AOR of 0.380) is also supported by Chen et al. (2022), who found that effective hypertension management often involves smoking cessation, reducing smoking rates in this group [43]. However, a study by Patel et al. (2023) found no significant association, suggesting that hypertension treatment alone may not always be a sufficient factor in reducing smoking [44].

### Study limitation

While the present study provides valuable insights into predictors of smoking behavior, there are several limitations to consider. Firstly, the cross-sectional nature of the study limits the ability to infer causality between the identified risk factors and smoking behavior. The reliance on self-reported data for smoking status and health conditions may also introduce reporting biases or inaccuracies. Additionally, the study’s sample may not be representative of the broader population, potentially limiting the generalizability of the findings. Factors such as socio-economic status, cultural influences, and access to healthcare services, which could also impact smoking behavior, were not comprehensively addressed in the analysis. These limitations should be taken into account when interpreting the results and designing future research.

### Study implication

The findings from this study underscore the importance of tailoring smoking cessation interventions to address specific risk factors such as elevated cardiovascular disease risk, high serum uric acid levels, and older age. Targeted programs that integrate cardiovascular health management and uric acid control could potentially enhance smoking cessation efforts. The observed link between hypertension and smoking suggests that individuals with hypertension might benefit from combined interventions that address both their hypertension and smoking habits[45]. By incorporating these insights, public health strategies can be more effectively designed to reduce smoking rates, particularly in populations with high-risk profiles. Additionally, the results highlight the need for continued research to explore how these factors interact and influence smoking behavior in different contexts.

## Conclusion

The study found that tobacco smoking is relatively uncommon among the population studied, with a very small percentage actively engaging in smoking. In addition, it identified elevated CVD risk, high serum uric acid levels, and older age as significant predictors of smoking. Conversely, having hypertension or being treated for it is associated with lower odds of smoking, possibly due to health interventions aimed at reducing smoking among those at higher risk. These findings emphasize the need for targeted interventions focusing on these specific risk factors when designing smoking cessation programs.

## Data Availability

All data related to this manuscript has been provided within this publication.

## Acknowledgements

The author extends their gratitude to all the study participants and the trained data collectors (Uwimana Jeanne D’Arc, Bibebityo Mariette, Aho Bantegeye Etienne) whose efforts were instrumental in making this study possible.

## Author Contributions

**Conceptualization:** Charles Nsanzabera.

**Data curation:** Charles Nsanzabera.

**Formal analysis:** Charles Nsanzabera, Jean claude Rukundo.

**Methodology:** Charles Nsanzabera, Leonard Ndayisenga.

**Writing – original draft:** Charles Nsanzabera.

**Writing – review & editing:** Charles Nsanzabera, Yusuf Said Mustafe, Jean claude Rukundo.

**Funding:** There was no financial support provided for conducting this study.

**Competing interests**: This study declared no competing interests.

## Availability of data and materials

Dataset was not share due to privacy of data.

## Notes

### Competing Interest Statement

The authors have declared no competing interest.

### Author Declarations

This study was carried out in accordance with the Declaration of Helsinki and was approved by the Rwanda National Ethical Committee (Reference number: 121/May/RNEC/2018). The ethical aspects, including confidentiality, voluntary participation, the option to withdraw, and the potential risks and benefits, were explained to all participants. Informed consent was obtained from each participant before the interviews and collection of biological samples took place.

